# Epigenetics of *PNLIPRP1* in human pancreas reveals a molecular path between type 2 diabetes and pancreatic cancer

**DOI:** 10.1101/2022.12.30.22284058

**Authors:** Lucas Maurin, Lorella Marselli, Lijiao Ning, Mathilde Boissel, Raphael Boutry, Mara Suleiman, Audrey Leloire, Vincent Pascat, Jared Maina, Bénédicte Toussaint, Souhila Amanzougarene, Alaa Badreddine, Mehdi Derhourhi, Inga Prokopenko, Anne Jörns, Sigurd Lenzen, François Pattou, Julie Kerr-Conte, Mickaël Canouil, Amélie Bonnefond, Piero Marchetti, Philippe Froguel, Amna Khamis

## Abstract

**Background:** Type 2 diabetes (T2D) increases the risk of pancreatic ductal adenocarcinoma (PDAC), which could be due to an epigenetic mechanism.

**Methods:** We explored the association between T2D and whole pancreas methylation in 141 individuals, of which 28 had T2D, using Illumina MethylationEPIC 850K BeadChip arrays. We performed downstream functional assessment in the rat acinar pancreas cell line AR42J. To further understand the role of our candidate gene in humans, we tested whether null variants were associated with T2D and related traits using the UK biobank.

**Results:** Methylation analysis identified one significant CpG associated with T2D: hypermethylation in an enhancer in *PNLIPRP1*, an acinar-specific gene. *PNLIPRP1* expression was decreased in T2D individuals. Using a rat acinar cell line, we 1/ confirmed decreased *Pnliprp1* in response to a diabetogenic treatment, and 2/ in *Pnliprp1* knockdown, an up-regulation of cholesterol biosynthesis, cell cycle down-regulation, decreased expression of acinar markers and increased expression of ductal markers pointing towards acinar-to-ductal metaplasia (ADM), a hallmark of PDAC initiation. Using exome data from UK Biobank, we show that rare *PNLIPRP1* null variants associated with increased glucose, BMI and LDL-cholesterol.

**Conclusions/interpretation:** We present evidence that an epigenetically-regulated gene associates with T2D risk, and might promote ADM and PDAC progression, opening new insights into early prevention of PDAC.

## 1. Introduction

Type 2 diabetes (T2D) is characterised by the inability of the pancreatic endocrine islets to secrete sufficient insulin in response to the body’s metabolic needs (1). In addition to this endocrine function, the pancreas has a distinct digestive exocrine function and its deterioration results in pancreatitis and/or in pancreatic ductal adenocarcinoma (PDAC), which remains the only type of frequent cancer with no major declines in mortality rates in the last years (2,3). Although the endocrine and exocrine pancreases have been studied as distinct entities, there is a strong interrelationship between the two pancreatic components: anatomically, the exocrine pancreas, which comprises > 98% of the pancreas (4), provides a structural scaffold and a favourable microenvironment for the development and optimal function of pancreatic islets, which account < 2% of the pancreas (5). On the other hand, blood circulation from the endocrine pancreas provides the delivery of high hormone concentrations from the islets to the exocrine tissue before entering the general circulation (5).

This crossover between endocrine and exocrine pancreas has also been demonstrated in epidemiological studies. Although there is no evidence to suggest a causative impact of T2D on pancreatitis, T2D patients have a 2.1 relative risk of developing PDAC (6). This is further supported by a recent Mendelian randomisation meta-analysis that suggested that T2D is causally associated with a higher PDAC risk (7). However, the molecular consequences of the systemic and local environment exposure to T2D states on the exocrine pancreas remain elusive.

Understanding DNA methylation dynamics can unravel fundamental mechanisms in response to deleterious environmental exposures that eventually may lead to severe diseases including cancer. Epigenome-wide association studies (EWAS) in pancreatic endocrine islets have identified methylated DNA sequences (CpGs) associated with T2D, implicating genes with roles in inflammation, glucose homeostasis, lipid metabolism and insulin resistance (8,9), which differ to those found by genetic studies (10). However, the potential impact of epigenetics to the exocrine pancreas, in the context of T2D, remains unexplored.

Therefore, we hypothesised that local pancreatic metabolic and hormonal changes associated with chronic hyperglycaemia exposure can lead to functional changes in the pancreatic exocrine tissue through methylome alteration. Identifying differentially methylated sites could pinpoint towards genes and molecular mechanisms linking T2D and pancreatic pre-cancer early stages. To identify these methylation changes, we performed an EWAS, in European organ donors with T2D and non-diabetic controls, to identify the epigenetic landscape associated with hyperglycaemia.

## 2. Methods

### 2.1. Epigenome wide association study in human samples

#### 2.1.1. Clinical characteristics

We selected 155 whole pancreas samples (147 brain-dead organ donors and 8 partial pancreatectomy patients), collected based on the IMIDIA consortium (11), according to sample availability. Of these samples, 32 had T2D, based on clinical characteristics and tests, according to the American Diabetes Association (ADA) guideline (ADA, 2019). The donors’ full clinical history and major laboratory parameters, such as blood glucose during the intensive care unit stay, were collected. Sample collection was followed by next-of-kin’s (for organ donors) or patient’s (for surgical cases) informed consent, and with the approval of the local ethics committees in Pisa and Hannover.

#### 2.1.2. DNA extraction, methylation arrays and statistical analysis

DNA was extracted from whole pancreas samples using the NucleoSpin Tissue kit (T740952.50; Mackerey-Nagel). Bisulphite conversion was performed in a total of 800 ng of DNA from our samples using the EZ DNA Methylation kit (5001; Zymo Research). To interrogate genome-wide DNA methylation status, our bisulphite converted DNA was subjected to Illumina’s 850K EPIC array. All DNA samples were genotyped using the Illumina Omni2.5M array and run on the Illumina iScan platform at the Imperial College London Centre (Hammersmith Campus, Imperial College London, UK). Methylation array data was imported using the *minfi* R package (12). Quality control (QC) steps removed CpG probes if they were: located on sex chromosomes or near single-nucleotide polymorphisms, cross-hybridising, non-cg or had a detection threshold p-value of less than 0.01. Samples with less than 99 % probes with a detection p-value lower than 0.01 were excluded. Probe-design biases and batch effects were normalised using R packages *Enmix* and *SVA* (13,14), respectively. Following QC, 746,912 probes remained. Samples were removed for having a call rate less than 99 % and two sex discordant samples were also removed. Following QC, 141 samples remained for further analysis (113 non-diabetic and 28 T2D individuals). Cohort statistics are summarised in Table 1. Population structure was evaluated by performing a principal component analysis (PCA) using a combined dataset of genomics data from 1,000 genomes and confirmed all samples were of European descent (Additional File 2: Figure S1).

**Table 1.** Organ donor summary statistics.

|  | **All**<br>**(n=141)** | **Non-diabetic**<br>**(n=113)** | **T2D**<br>**(n=28)** | **P-value** |
| --- | --- | --- | --- | --- |
| **Sex (% male)** | 72 (51.1%) | 52 (46.0%) | 20 (71.4%) | 0.028 |
| **Age (SD)** | 65.6 (14.9) | 63.7 (15.6) | 73.4 (7.58) | 0.004 |
| **BMI (SD)** | 25.6 (3.56) | 25.3 (3.46) | 26.6 (3.88) | 0.102 |
| **Glycaemia (SD)** | 9.05 (2.83) | 8.33 (2.25) | 11.9 (3.13) | 0.0001 |
|  | (n=124) | (n=99) | (n=25) |  |
| *Standard deviation |  |  |  |  |

A linear regression model was applied to associate T2D with CpG methylation level at a given probe. We corrected for age and sex. We performed a linear model including BMI as a co-variate, and found that BMI did not improve the model (Additional File 1: Table S1). Whole pancreatic tissue samples were expected to include a variety of cell types and thereby be a potential confounding effect on DNA methylation. Therefore, cell composition was estimated using the R package *RefFreeEWAS* (15). Multiple testing using a strict Bonferroni correction was applied and CpGs with an adjusted p-value < 0.05 were considered significant. The EWAS model is detailed below:

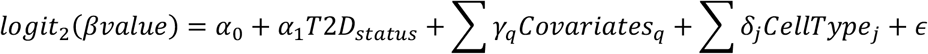

with covariates as age and sex. The study design is summarised in Figure 1.

**Figure 1.**
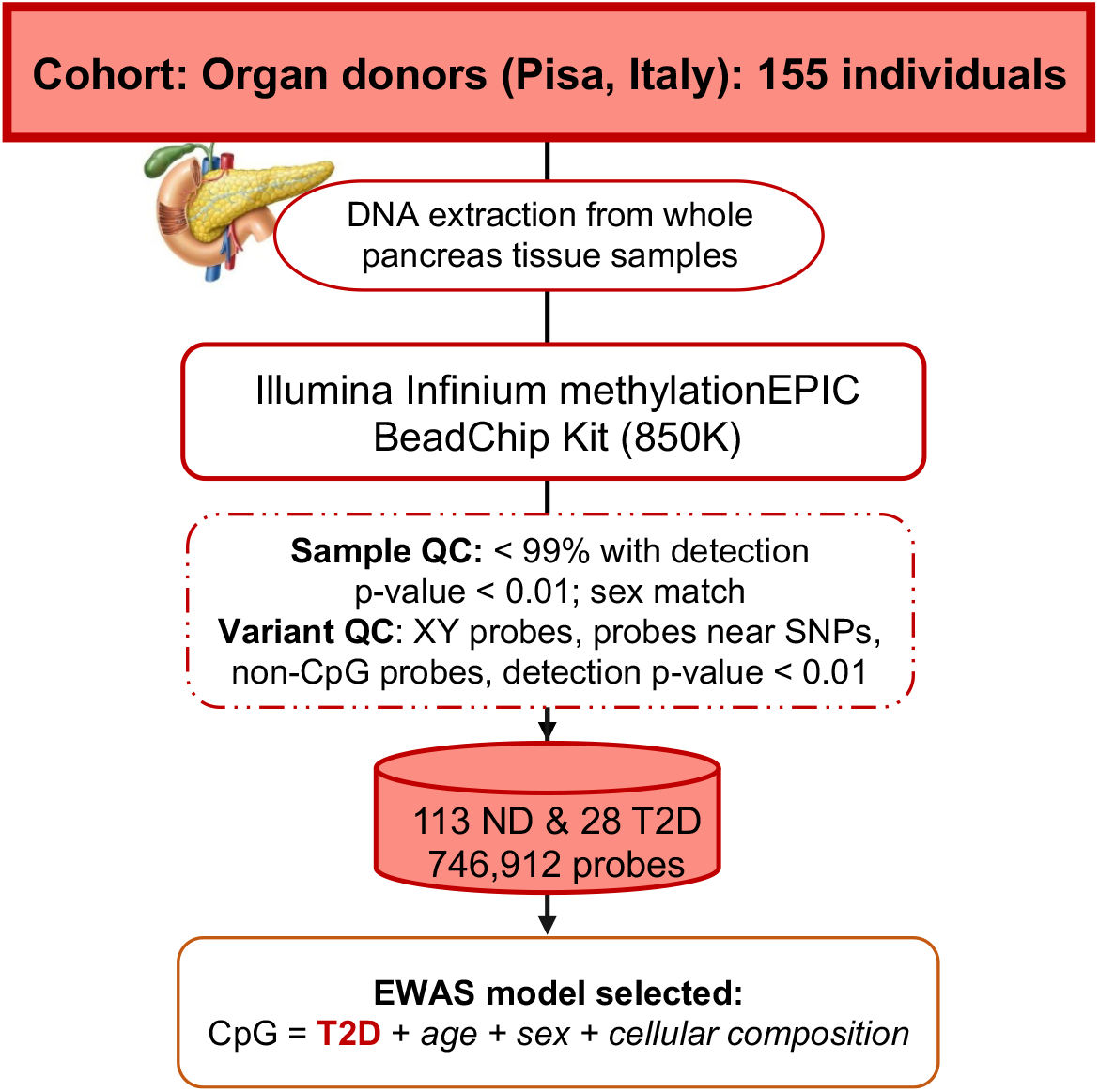
Overview of method used to identify methylation sites associated with T2D. DNA was obtained from organ donor pancreases and methylation was performed. Following QC, several epigenome wide association studies were performed and the best model was chosen to identify methylation changes associated with T2D. ND: non-diabetic controls.

#### 2.1.3. Differentially methylated regions (DMR) and enhancers

To identify DMRs, the Bioconductor package DMRcate was used (Peters et al., 2015). DMRcate determines regions based on EWAS results. A CpG with p-value < 0.05 was considered significant and we defined a DMR as a region with at least 2 CpGs within the Gaussian kernel bandwidth (lambda) equals to 1 kb. To determine if specific CpGs and DMRs were situated within regulatory regions, we consulted the Genehancer (Fishilevich et al; 2017) and dbSUPER (Khan et al; 2016) database.

### 2.2. RNA expression of *PNLIPRP1* and neighbouring genes in cohort

Prior to processing, the samples were stored in OCT at −80°C. Ten samples were processed (five controls and five T2D individuals), matched for age, sex and BMI (Additional File 1: Table S3). Detailed method is in Additional File 1: Supplementary Methods.

### 2.3. Immunohistochemistry

protein expression of PNLIPRP1 in human tissues Human pancreatic tissue sections, from control and T2D individuals, used in this study were donated by Inserm UMR1190 unit (University of Lille, France). We excluded any fibrotic process or pathological alterations in the pancreas tissues. Immunohistochemistry method is detailed in the supplementary.

### 2.4. Functional characterisation

#### 2.4.1. Cell line, RNA extraction and qPCR

To characterise the role of *PNLIPRP1* and neighbouring genes, all *in vitro* assays were performed using the rat AR42J acinar cell line (CRL-1492; ATCC). For growth, cells were incubated at 37°C and 5% CO_2_ with RPMI 1640 Glutamax medium (61870044; Gibco), supplemented with 10% FBS (26140079; Gibco), 0.01% penicillin/streptomycin (P/S) (15410-122; Life Technologies). Medium was changed every 48 hours.

#### 2.4.2. High glucose and insulin treatment

AR42J cells (2×10^5^ per well) were plated in 6 well plates (4413; Beckton Dickinson) and grown to confluence. Upon reaching confluence, the medium was removed and replaced with a 0.1% FBS (26140079; Gibco) version of the medium previously described along, with 14 µL of 1M glucose or 5 µL of 20 µM insulin, or both, for 72 hours. Following incubation, cells were washed with 1 x PBS and then harvested for RT-qPCR. The experiment was performed in three biological replicates. Gene expression differences were tested using a two-way ANOVA were performed using GraphPad Prism (GraphPad software Inc).

#### 2.4.3. siRNA knockdown

To perform a *Pnliprp1* KD in AR42J, the cells were transfected in suspension using the AR42J Transfection Reagent kit (1181; Altogen) along with *Pnliprp1* siRNA (M-099515-01-0010; Dharmacon) or non-targeting control siRNA (D-001810-10-20; Dharmacon) following the manufacturer’s instructions. Following transfection, RNA and protein from siRNA treated AR42J cells were harvested after 48 or 72 hours. This experiment was performed in four biological replicates. To confirm the down-regulation of *PNLIPRP1*, we performed a qPCR for *Pnliprp1* of all the biological replicates and tested using a two-tailed t-test in Graphpad Prism. In addition, we extracted protein from *Pnliprp1* KD and performed western blotting of Pnliprp1 and beta-actin to confirm the down-regulation at the protein level. MTS proliferation (197010; Acbam) and Cholesterol Ester-Glo Assay (J3190; Promega) methods performed in *PNLIPRP1* KD is detailed in the supplementary methods.

#### 2.4.4. RNA Sequencing

RNA extracted from *Pnliprp1* KD in AR42J cells from four biological replicates were subjected to RNA sequencing. Library preparation was performed using the KAPA mRNA HyperPrep Preparation Kit (Roche) following the manufacturer’s instructions. Sequencing was performed with the NovaSeq 6000 (Illumina). Mean sequencing depth was of 100 million 100 bp paired-end reads per sample. Illumina Raw data were demultiplexed using bcl2fastq v2.20.0.422 (Illumina) and adapters trimming step has been executed using cutadapt version 3.2. Mapping was done using STAR version 2.7.1a with Rattus_norvegicus.Rnor_6 genome. Raw and normalised counting steps were done using RSEM v1.3.0 using a GTF from Ensembl version 102, and Ensembl v.102 for genes names annotations. The differential expression analysis was performed using DeSeq2.

##### 2.4.4.1. Pathway analysis of RNA sequencing data

Over-representation analysis (ORA) used to determine whether significantly expressed genes (p<0.05) are overrepresented in a particular pathway (16). The adjusted p-value cut-off for reported enriched terms was set to 0.05. In addition, Ingenuity Pathway Analysis (IPA) (Qiagen) was used for pathway analysis and EnrichR (Kuleshov et al, 2016) were used to identify enriched gene ontology terms and pathways in several databases.

#### 2.4.5. UK Biobank to identify rare and common variant associations

Exome sequencing data from up to 191K participants in the UK biobank was used to identify rare null variants (MAF < 1%) in *PNLIPRP1*. This research is part of UK Biobank research application #67575. We tested association of null variants for BMI, glucose and lipid traits using the MiST method (17), which tests rare variants in a single cluster (at the gene scale). This method provides a score *π*which represents the mean effect of the cluster and a score *τ*which represents the heterogeneous effect of the cluster. The overall p-value tests the association between the set of variants and the trait of interest.

For each trait, we adjusted for relevant co-variates (detailed in Additional File 1: Table S5). We considered a trait as significant if the p-value associated with the direct burden effect of the cluster was significant (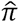 p-value i.e. P_val < 0.05) and the direction of effect for the variants was consistent, revealed by the absence of heterogeneity (tau p-value *i*.*e*., P.value.S.tau > 0.05).

#### 2.4.6. *PNLIPRP1* DNA variant association analyses for pancreatic cancer in UK Biobank and for T2D in publicly available summary statistics

UK biobank analyses. We performed the GWAS for pancreatic cancer in the UK biobank using a linear mixed model (LMM) to test for association implemented in the software tool BOLT-LMM (18). For association analysis of common variants (MAF > 0.01) we applied the following quality control criteria – imputation score > 0.4, Hardy-Weinberg Equilibrium (HWE) P-value > 1×10^−6^, and SNP variant missingness rate < 0.015. We included age, sex, genotyping array and six principal components (PCs) as covariates in the LMM.

In the *PNLIPRP1* gene region, we verified the associations with common and rare variant SNPs in two publicly available GWAS summary statistics datasets for T2D and PDAC, respectively. For analysis, we evaluated a DNA region window of five times the *PNLIPRP1* gene region. We used the latest T2D GWAS from Mahajan et al. (19), including 74,124 T2D cases and 824,006 controls of European ancestry.

## 3. Results

### 3.1. Epigenome-wide association study reveals candidate gene in T2D

In order to identify potential epigenetic modifications associated with T2D in the exocrine pancreas, we measured methylation levels in the whole pancreas tissue from 155 individuals. Following QC, 141 individuals, of which 28 had T2D, were available. We performed a linear regression, adjusted for age, sex and cellular composition,f to identify methylation changes associated with T2D status (Figure 1). Following multiple testing using Bonferroni correction, we identified a single significant association between increased T2D risk and a hypermethylation at the cg15549216 probe (adjusted p-value = 0.02) (Figure 2a), located in the gene body of *PNLIPRP1*, encoding pancreatic lipase related protein 1. This CpG was 11.4% more methylated in individuals with T2D, compared to controls (Figure 2b; estimate = 0.6; standard error = 0.1).

**Figure 2.**
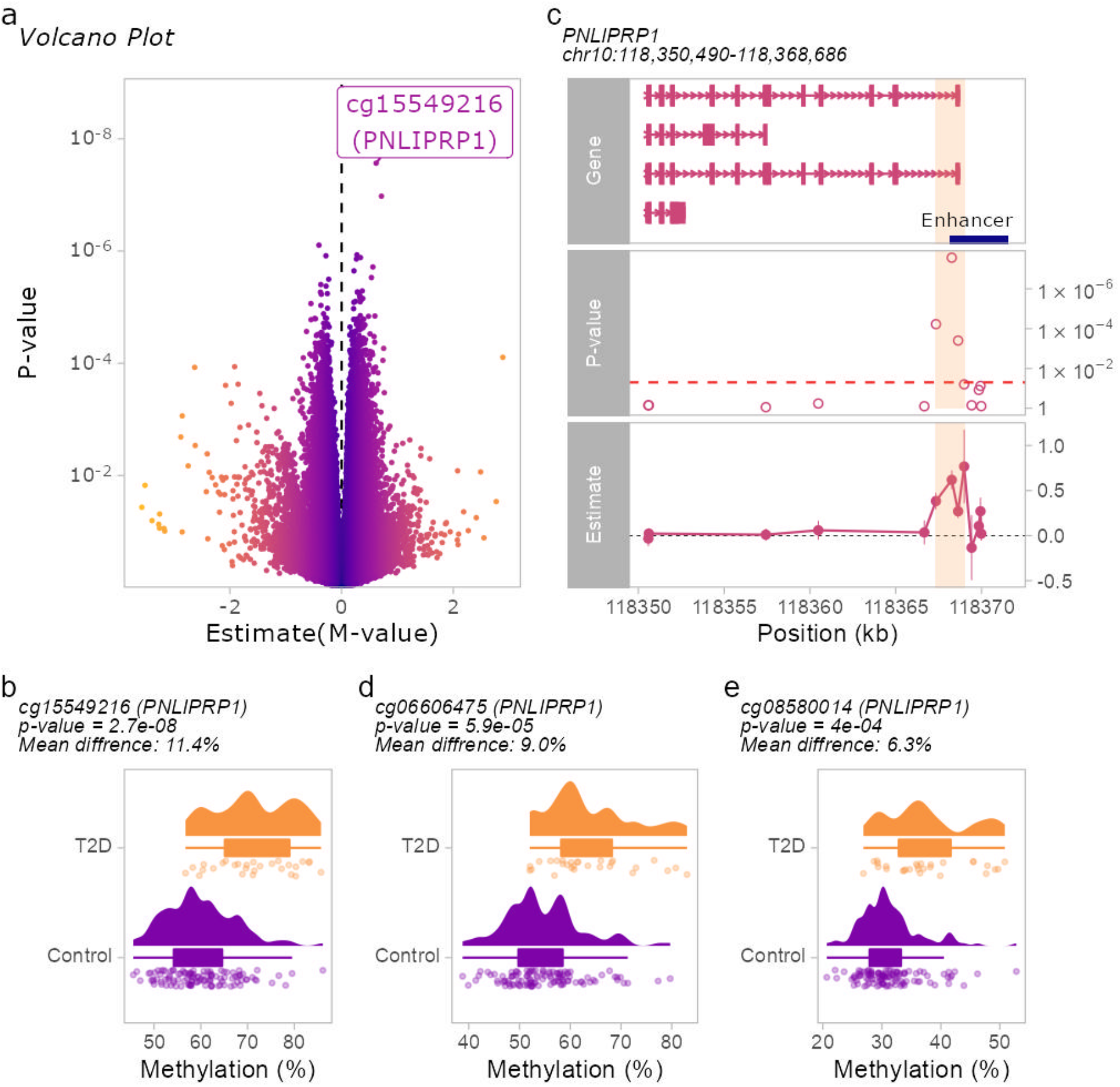
Epigenome-wide association study in whole pancreas in T2D. a) Volcano plot depicting the differentially methylated CpGs associated with T2D. The labelled CpGs have a Bonferroni-adjusted p-value < 0.05. b) Boxplot showing the density of cg15549216 methylation levels in T2D individuals and non-diabetic controls, for each individual. The X-axis represents the % methylation beta-value. c) A representation of the *PNLIPRP1* gene, enhancer region, the estimate and p-values of all CpGs in the *PNLIPRP1* gene, and the DMR is shaded in red. d-e) Boxplot showing the density of cg06606475 and cg08580014 methylation levels, respectively, in T2D individuals and non-diabetic controls, for each individual.

To identify whether this CpG was a part of a region of dysmethylated CpGs, we performed a differentially methylated region (DMR) analysis to find regions with at least two or more CpGs associated with T2D within a window of 1 kb. We found three CpGs in this region, including the aforementioned cg15549216 probe, in addition to two flanking CpGs, cg06606475 and cg08580014 (Figure 2c). The cg06606475 probe is located in the gene body, 921 base pairs (bp) upstream of the cg15549216 probe (Figure 2d; mean difference = 9.0 %; p = 5.9 × 10^−5^; estimate = 0.38; standard error = 0.09), and cg08580014, located in the 3’UTR of *PNLIPRP1*, is 370 bp downstream of the cg15549216 probe (Figure 2e; mean difference = 6.3 %; p = 4.0 × 10^−4^; estimate = 0.27; standard error = 0.07). These CpGs were individually nominally associated with T2D in our EWAS and consistent in direction of effect. To identify whether the cg15549216 CpG was also associated with glucose traits, we performed a linear regression of CpG and glucose levels in our individuals and found that the CpG was positively correlated with glucose levels (p = 0.00017; correlation rho = 0.33; Additional File 2: Figure S2).

*PNLIPRP1* is located between two related family member genes: *PNLIP* (pancreatic lipase) and *PNLIPRP2* (pancreatic lipase related protein 2) (Additional File 2: Figure S3). Therefore, we hypothesised that this region is a regulatory region involved in the regulation of pancreatic lipase genes. To explore the potential regulatory and functional significance of this locus, we checked already published open chromatin resources. The Genehancer and dbSUPER enhancer databases showed that ChIP-Seq data from human whole pancreas tissue samples (GSM1013172) in and surrounding the cg15549216 probe lies in a 12 kb stretch of the only known enhancer of *PNLIPRP1* (Figure 2c).

### 3.2. Correlation of cg15549216 CpG with nearby gene expression

To determine whether the expression of *PNLIPRP1, PNLIP* and *PNLIPRP2* were dysregulated in T2D, we performed gene expression quantification of the respective genes. To do this, we selected a subset of five individuals with T2D and six non-diabetic controls, matched for age, sex and body mass index (BMI) (Additional File 1: Table S3), and extracted RNA from whole pancreas tissue. We found that the expression of *PNLIPRP1* was down-regulated in T2D (53%, p = 0.0109), but was unchanged for *PNLIP* and *PNLIPRP2* (Additional File 2: Figure S4; p = 0.65 and p = 0.059 respectively).

To determine whether methylation differences had a functional impact on *PNLIPRP1*, we performed a linear regression of methylation levels and the deltaCT, *i*.*e*., the absolute difference between gene expression and the housekeeping gene: a higher deltaCT indicates a lower expression of the gene, and *vice versa*. We found that cg15549216 significantly increased methylation correlated with decreased *PNLIPRP1* gene expression (Figure 3b; p = 0.042; R^2^ = 0.35).

**Figure 3.**
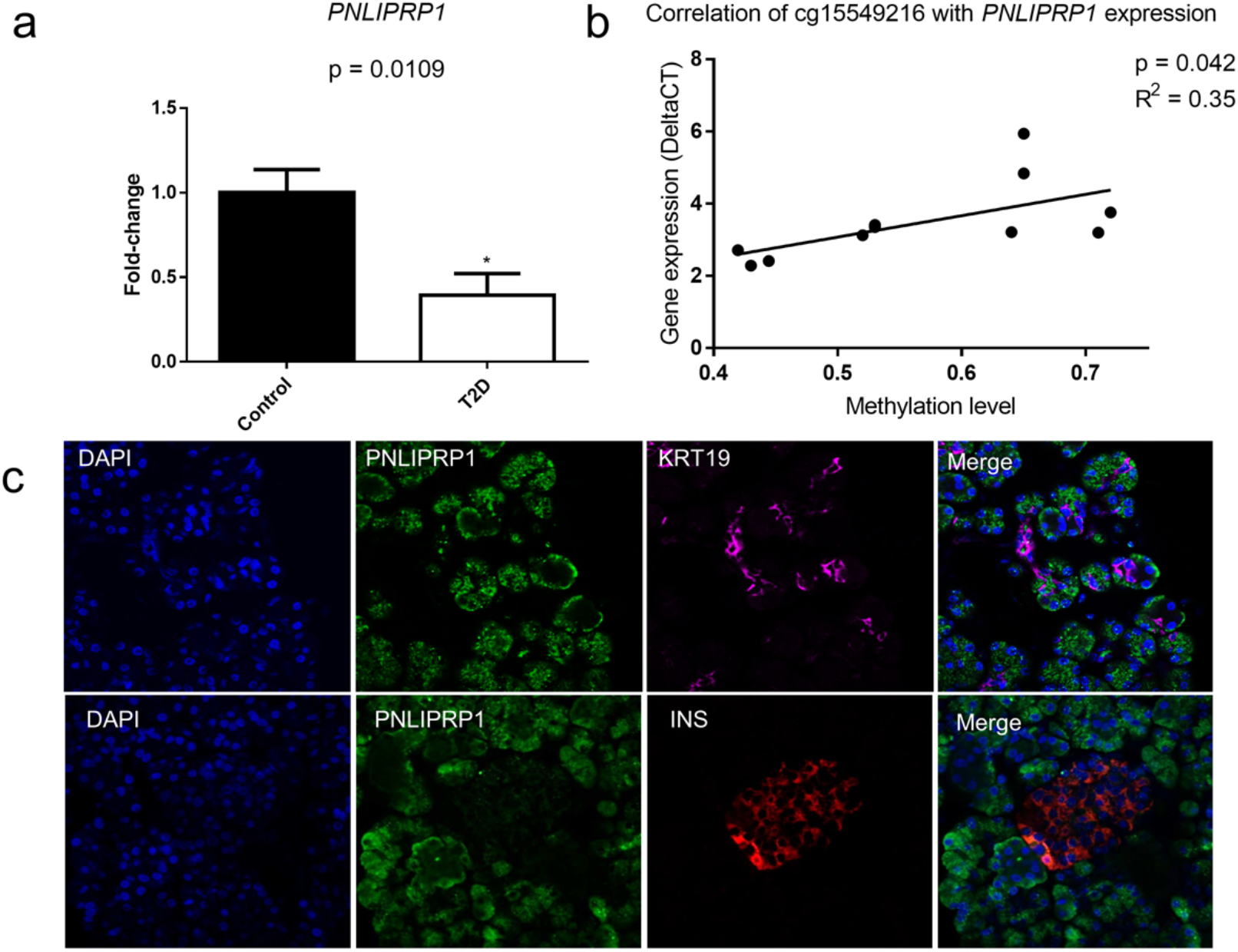
Expression of PNLIPRP1 at the RNA and protein level. a) RNA expression of *PNLIPRP1* in a subset of 11 individuals (5 with T2D *vs*. 6 non-diabetic controls), matched for age, sex and BMI. T-tests were performed to test for differences between T2D and non-diabetic samples for each target gene. Error bars represent the standard error. b) Correlation between the cg15549216 probe and the RNA expression of the *PNLIPRP1* gene determined by qPCR. DeltaCT represents difference in expression between the respective gene and the housekeeping gene *RPLP0*. Higher DeltaCT values indicate a reduction in gene expression. c) Immunofluorescence of healthy human whole pancreas tissue samples stained for PNLIPRP1 and KRT19, a ductal marker (top), and INS (insulin – a marker for pancreatic islets). Nuclei were stained with DAPI.

To identify target cell types of *PNLIPRP1* dysregulation, we examined the expression of *PNLIPRP1* in the GTEx database (20), which includes tissue expression of 53 human tissues. It showed that the *PNLIPRP1* was exclusively expressed in the whole pancreas (Additional File 2: Figure S5). As the pancreas mainly includes acinar, ductal and pancreatic islets, we analysed several single-cell pancreas datasets (21,22) and found that *PNLIPRP1* is highly and solely expressed in acinar cells, but not in ductal cells of the exocrine pancreas, and not in pancreatic islet alpha, beta, delta and pancreatic polypeptide cells (Additional File 1: Table S6). These data point towards an acinar-specific expression of *PNLIPRP1*. To confirm at the protein level the acinar expression of *PNLIPRP1*, we performed immunofluorescence in human pancreatic tissue stained for PNLIPRP1, and also for KRT19, a marker of ductal cells, and for insulin to mark pancreatic islets. It revealed that the protein PNLIPRP1 was not expressed in pancreatic islets or ductal cells, but exclusively in acinar cells (Figure 4c).

**Figure 4.**
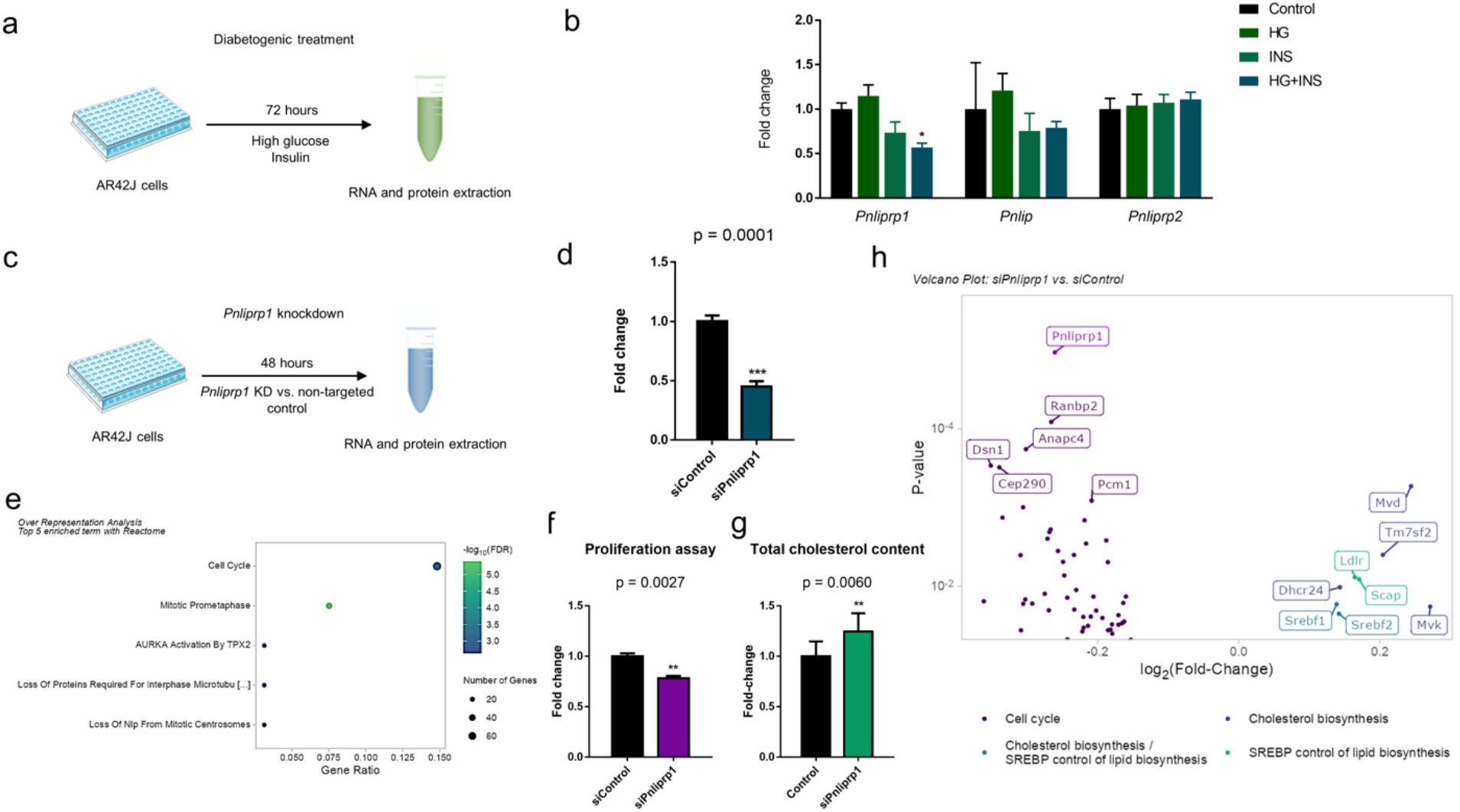
Functional characterisation *Pnliprp1* in Ar42J rat cell line. a) AR42J were treated with either glucose, insulin, or both, for 72 hours, and RNA and protein were extracted. b) RNA expression of *Pnliprp1* and neighbouring related genes, *Pnlip* and *Pnliprp2*, was determined by qPCR. A two-way ANOVA was performed to assess differences in expression between the control and different treatments and target genes. Error bars represents the standard error. ** = p< 0.01. c) Method used to perform *Pnliprp1* KD in AR42J. d) Confirmation of *Pnliprp1* KD in four biological replicates using qPCR and tested using a t-test. e) Overrepresentation analysis of the top five most dysregulated pathways in *Pnliprp1* KD using Reactome database. f) MTS proliferation assay in AR42J in *Pnliprp1* KD, performed in three biological replicates after 72 hours of siRNA treatment. A t-test was performed to assess differences in proliferation between the siPnliprp1 and non-targeting control. Error bars represents the standard error. g) Total cholesterol measured following 48 hours of *Pnliprp1* KD, performed in three biological replicates. Error bars represents the standard error. h) Volcano plot of RNA sequencing results, depicting *Pnliprp1* and genes from the most dysregulated up and down-regulated pathways based on EnrichR database.

### 3.3. Functional characterisation of *Pnliprp1* in rat acinar cell line AR42J

#### 3.3.1. Reduction of *Pnliprp1* in response to diabetogenic treatment

Based on the acinar-specific expression pattern of PNLIPRP1, we explored the functional role of *PNLIPRP1* in the rat acinar cell line AR42J. To address whether T2D induces a dysregulation of *PNLIPRP1* in our model, we tested whether a diabetogenic environment (*i*.*e*., high glucose and insulin) induced a change in *Pnliprp1* expression (Figure 4a). We first confirmed that the AR42J cells are responsive to insulin treatment by analysing phosphorylated Akt, a measure of insulin signalling (Additional File 2: Figure S6). We found that the treatment of high glucose or insulin alone did not change *Pnliprp1* expression, however, the combination of both treatments induced a 35% decrease in *Pnliprp1* expression (Figure 4b). This effect was not observed in nearby *Pnlip* and *Pnliprp2* genes (Figure 4b), confirming our human data, which showed that T2D induces a dysregulation specific to *PNLIPRP1*.

#### 3.3.2. *Pnliprp1* KD in acinar AR42J cells induced a down-regulation of cell cycle and up-regulation of cholesterol biosynthesis genes

To investigate the downstream consequence of *Pnliprp1* dysregulation, we performed a knockdown (KD) of *Pnliprp1* in AR42J cell line (Figure 4c). We verified that *Pnliprp1* KD induced a 60% decreased gene expression of *Pnliprp1* (Figure 4d) and 34% decrease at the protein level (Additional File 2: Figure S7). We then performed RNA sequencing to identify the transcriptomic changes in response to *Pnliprp1* KD. We confirmed that *Pnliprp1* was one of the most significant down-regulated genes (FDR = 0.02), and the expression of *Pnlip* and *Pnliprp2* was unchanged (p > 0.05). Using an FDR significance cut-off of 0.05, in addition to *Pnliprp1*, we identified 10 other dysregulated genes: *Rbm39, Clk1, Cpne1, Dld, Sf3b1* involved in cancer, *Cyp8b1 in* lipid metabolism and *Pnisr, Zfp386, Luc7l3, Gpcpd1* genes of unknown roles (Additional File 1: Table S7). We explored potential dysregulated pathways by focusing on the 1,024 genes with an unadjusted p-value < 0.05 (Additional File 1: Table S7) and utilised several tools to identify pathways dysregulated in our KD model. Using an over-representation analysis (ORA) based on the Reactome database, we found that the “cell cycle” was the most dysregulated pathway (FDR < 0.05) (Figure 4e). This was further confirmed using IPA (Additional File 2: Figure S8), where the top down-regulated pathway was the ATM signalling pathway, which has a primary role in activating cell cycle checkpoints (23), with a decrease in the expression of key genes *Atm, Rad50, Brca1, Atf2, Hp1, Cdk1* and *Smc* (Additional File 2: Figure S9).

In order to confirm and quantify this down-regulation of the cell cycle, we performed in the *Pnliprp1* KD an MTS proliferation assay, used to assess and measure cellular proliferation. As expected, it resulted in a 22% reduction in cell proliferation in the *Pnliprp1* KD, compared to non-targeting controls (p = 0.0027) (Figure 4f).

To determine if the transcriptome overrepresentation of the cell cycle may have masked other relevant pathways, we explored the up and down regulated pathways separately using EnrichR, which includes several pathway analysis tools. We confirmed that the top pathway for the down-regulated genes (587 genes) was the cell cycle pathway (adjusted p < 0.0001; Table 2; Additional File 1: Table S8). In contrast, we found that the most up-regulated pathways (437 genes) were “cholesterol biosynthesis” and “SREBP control of lipid biosynthesis” (adjusted p < 0.0001; Table 2; Additional File 1: Table S9), due to the up-regulation of key genes involved in maintaining cellular cholesterol homeostasis. It included the crucial transcription factors, sterol regulatory element binding proteins 1 and 2 (*Srebf1* and *Srebf2*), in addition to key enzymes mevalonate phosphate decarboxylase (*mvp*), mevalonate kinase (*mvk*), the sterol reductase *Tm7sf2* and 24-Dehydrocholesterol Reductase (*Dhcr24*), all of which were up-regulated in our *Pnliprp1* KD (Figure 4h). We confirmed that total cholesterol content in increased 29 % following *Pnliprp1* KD (p = 0.0017; Figure 4g). Taken together, we demonstrated that the invalidation of PNLIPRP1 dysregulates in inverse direction cholesterol metabolism and cell cycle regulation.

**Table 2.** Enriched pathways following the KD of *Pnliprp1* in AR42J

| **Source** | **Pathway** | **Adjusted p-value** | **Odds ratio** |
| --- | --- | --- | --- |
| **Upregulated pathways** |  |  |  |
| Reactome | Cholesterol biosynthesis | $4.3 \times 10^{-3}$ | 15.12 |
| BioCarta | SREBP control of lipid biosynthesis | $4.3 \times 10^{-3}$ | 47.17 |
| KEGG | Metabolism | $4.3 \times 10^{-3}$ | 1.89 |
| KEGG | Arginine and proline metabolism | $4.3 \times 10^{-3}$ | 7.91 |
| Reactome | Lipid and lipoprotein metabolism | $4.3 \times 10^{-3}$ | 2.61 |
| **Downregulated pathways** |  |  |  |
| Reactome | Cell cycle | $2.21 \times 10^{-12}$ | 4.29 |
| | Messenger RNA processing | $1.69 \times 10^{-11}$ | 6.25 |
| Reactome | Capped intron-containing pre-mRNA processing | $2.93 \times 10^{-9}$ | 6.86 |
| | DNA replication | $2.93 \times 10^{-9}$ | 5.39 |
| | Messenger RNA splicing: major pathway | $1.32 \times 10^{-7}$ | 9.59 |

#### 3.3.3. *Pnliprp1* KD induced acinar-to-ductal metaplasia

Alterations in proliferation and in cellular identity are often linked, and indeed our RNA sequencing data revealed dysregulation in several acinar markers in *Pnliprp1* KD, which were subsequently confirmed by qPCR. Four acinar markers were down-regulated: *Prss1* (p = 0.0066), *Amy2* (p = 0.0029), *Cpa2* (p = 0.035). *Ctrl* was down-regulated in three of the four replicates (p = 0.09) (Figure 5a); in contrast, two ductal markers were up-regulated: *Krt19* (p = 0.0001) and *Hnf1b* (p = 0.020) (figure 5b). Taken together, these results suggest that *PNLIPRP1* invalidation may induce acinar-to-ductal cellular trans-differentiation.

**Figure 5.**
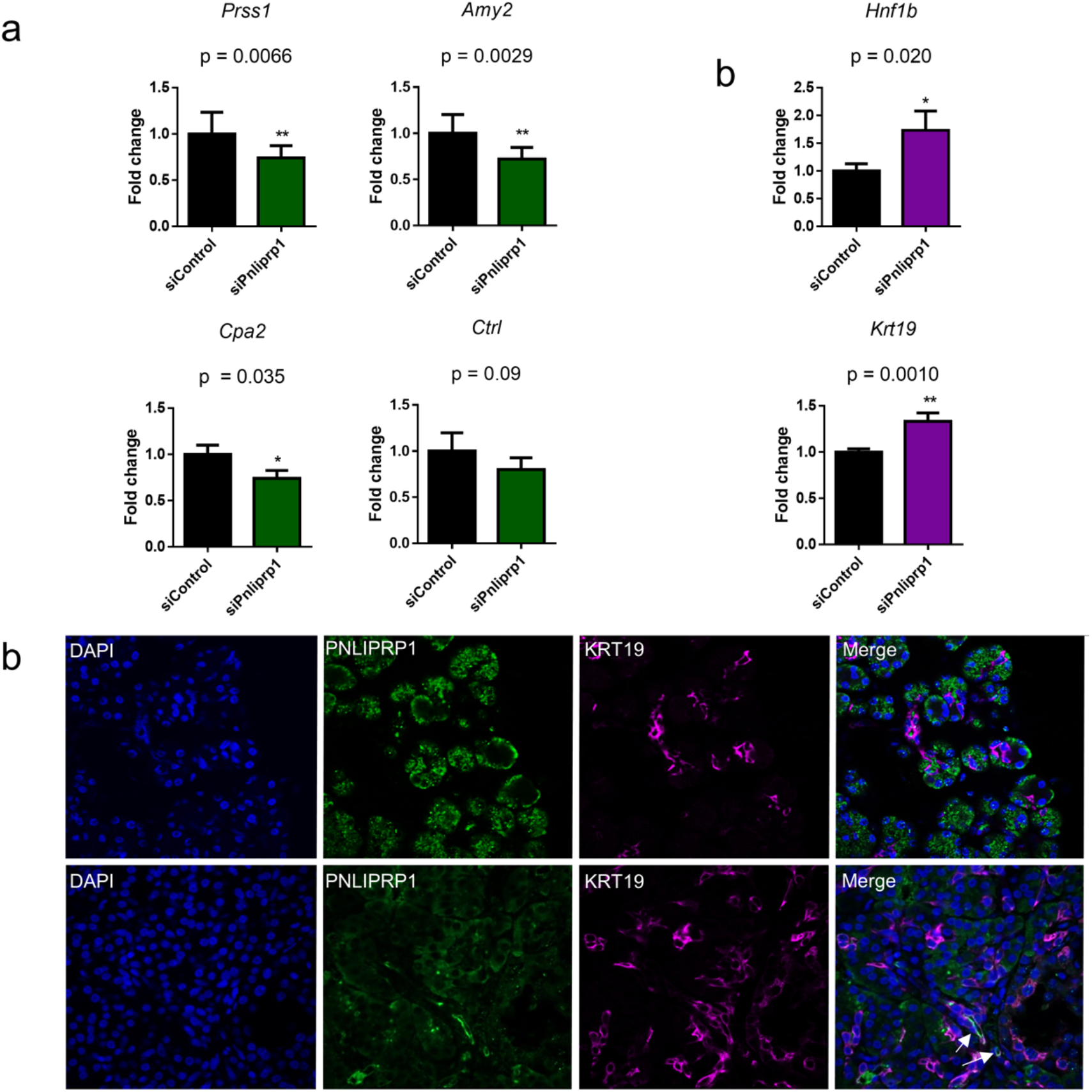
Acinar-to-ductal metaplasia observed in AR42J and human tissue. a-b) Validation of acinar and ductal marker dysregulation seen in transcriptomics *Pnliprp1* KD, confirmed using qPCR. T-tests were performed between each target gene and the untargeted control. Error bars represent one standard error. n=4; * = p < 0.05; ** = p < 0.01. b) Immunofluorescence of human whole pancreas tissue from non-diabetic (top) and T2D individuals (bottom). The tissue samples were stained for PNLIPRP1 and KRT19, a ductal cell marker. Nuclei were stained with DAPI. White arrows in the T2D merge image depict co-expression of PNLIPRP1 and KRT19 cells.

To assess whether this trans-differentiation occurs in human tissues from patients with T2D, we stained pancreatic tissue from patients with T2D and from non-diabetic controls with both PNLIPRP1 and ductal marker KRT19. We observed a higher KRT19 protein expression in individuals with T2D (Figure 5c). Intriguingly, we also observed co-expression of both PNLIPRP1 and KRT19 in some cells from patients with T2D, which was not seen in non-diabetic controls.

#### 3.3.4. Association of *PNLIPRP1* variants with T2D and related traits

##### Rare null variants in PNLIPRP1

In order to test whether rare variants (minor allele frequency [MAF] < 1%) in the *PNLIPRP1* gene were associated with T2D and related metabolic traits, we used exome sequencing data from up to 191K participants in the UK biobank. We identified 45 null variants (*i*.*e*. nonsense, frameshift, canonical ±1 or 2 splice sites). We considered T2D and associated metabolic traits, including BMI, glucose and lipid traits (Table 3). We found that *PNLIPRP1* null variants were associated with increased glucose levels (p = 1.1× 10^−3^; effect size = 0.13; SE = 0.04), and this association remained without BMI adjustment (p = 4.6× 10^−4^; effect size = 0.16; standard error = 0.05). In addition, we found that *PNLIPRP1* null variants were associated with log BMI (p = 3.1 × 10^−4^; effect size = 0.04; standard error = 0.01) and waist circumference (p = 7.6× 10^−3^; effect size = 2.19; standard error = 0.82). We also found that *PNLIPRP1* rare variants were significantly associated with low density lipoprotein cholesterol (LDL-C; p = 0.03; effect size = 0.10; standard error = 0.05). However, null variants were not associated with T2D (p = 0.48), obesity (p = 0.29), or log triglyceride levels (p = 0.88) (Additional File 1: Table S5). Altogether, our data confirms that null *PNLIPRP1* variants are associated with increased glucose levels, weight and LDL-cholesterol. In this respect, our own recent data using Mendelian randomisation suggests that BMI, and particularly visceral fat, might be the causative driver of the link between T2D and PDAC (24).

**Table 3.** Association of null *PNLIPRP1* variants with T2D associated traits

| Trait | Sample size | pi p-value | tau p-value | Effect size | Standard error | Rare variants |
| --- | --- | --- | --- | --- | --- | --- |
| Glucose (mmol/l) | 159764 | $1.1 \times 10^{-3}$ | 0.13 | 0.13 | 0.04 | 38 |
| Log BMI (kg/m<sup>2</sup>) | 187727 | $3.1 \times 10^{-4}$ | 0.07 | 0.04 | 0.01 | 40 |
| Waist circumference (cm) | 190751 | $7.6 \times 10^{-3}$ | 0.19 | 2.19 | 0.82 | 41 |
| LDL cholesterol (mmol/l) | 169625 | 0.03 | 0.20 | 0.10 | 0.05 | 38 |

##### Common variants in PNLIPRP1

We used the Type 2 Diabetes Knowledge Portal, which analyses genetic association results and aggregates genome wide association studies (GWAS) to identify single nucleotide polymorphism (SNPs; MAF > 1%) associated with T2D and related traits (Additional File 2: Figure S10). We found that 27 SNPs located in *PNLIPRP1* were associated with LDL-C, high density lipoprotein (HDL), and cholesterol levels following multiple testing (p ≤ 2.5 × 10^−6^) (Additional File 1: Table S10).

In addition, using latest T2D GWAS (N=898,130; 74,124 cases; 824,006 controls) and our in-built pancreatic cancer GWAS from UK Biobank (N=458,890: 1,425 cases and 457,465 controls), we tested for genetic association of SNPs located within the *PNLIPRP1* with T2D and PDAC and did not find any significantly associated SNPs for both traits (Additional File 2: Figure S11 and S12). Altogether, genetic studies suggest that *PNLIPRP1* variants can be causally associated with glucose and lipid metabolism but not with T2D risk.

Altogether, genetic studies suggest that *PNLIPRP1* variants can be causally associated with glucose and lipid metabolism but not with T2D risk.

## 4. Discussion

To our knowledge, we have performed the first EWAS study to explore the epigenetic associations with T2D in the whole pancreas, which comprises > 98% exocrine pancreas, a tissue, which usually is not directly involved in T2D early development, but in one of its most severe complication. The cg15549216 CpG hypermethylation, located in the only known enhancer of *PNLIPRP1*, and more generally the region in its vicinity, strongly associated with T2D status, and was inversely correlated with the expression of this gene.

We believe that *PNLIPRP1* is a strong candidate to (epi)genetically link T2D (and obesity) with the chain of events potentially leading to further PDAC for several reasons: 1/ PNLIPRP1 is highly and solely expressed in the acinar cells and 2/ The exposure of rat acinar cell lines to diabetogenic milieu downregulated *Pnliprp1*. In addition, *Pnliprp1* KO mice displayed mild hyperglycaemia and impaired insulin sensitivity and a high fat diet induced an exacerbation of insulin resistance, glucose intolerance and increased fat mass (25). This corresponds to our human genetic data, suggesting that *PNLIPRP1* variation were associated with metabolic abnormalities and with body corpulence (both BMI and waist circumference). Taken together, our study demonstrates that both nature (genetic variation) and nurture (through epigenetic marks) may synergistically contribute to the exacerbation of *PNLIPRP1* dysregulation in acinar cells, creating a deleterious vicious cycle, contributing to changes in the fate of some cells of the exocrine pancreas.

Our functional genomics analyses indeed suggested that diminished *PNLIPRP1* expression associated with changes in cell cycle regulation and in acinar identity. This de-differentiation and cell cycle arrest are two hallmarks of acinar-to-ductal metaplasia (ADM) (Wang et al., 2019). Although in principle this ADM is reversible, during sustained stress, these cells are unable to revert to a differentiated state, making a particularly favourable environment for *Kras* mutations, Tert expression and TGF-beta signalling, molecular events that strongly associate with the progression of ADM to PDAC (27–30). In this respect, a recent meta-analysis of publicly available PDAC and matched adjacent tissue, found that *PNLIPRP1* was the third most down-regulated gene in PDAC (31). Additionally, a recent single cell study in PDAC tissues in mice, found that *Pnliprp1* was down-regulated in early metaplastic PDAC and further exacerbated in late stage metaplasia (32), demonstrating a potential role of *PNLIPRP1* in early ADM, but also in sustained progression into PDAC.

Additionally, several studies have shown that cholesterol biosynthesis and uptake pathways are highly enriched in PDAC, including key genes involved in the process, such as the low-density lipoprotein (LDLR) and sterol regulatory element-binding protein 1 (SREBP1), which have been proposed as prognostic markers of PDAC (33,34). In addition, randomised control trials and retrospective studies have shown that statins, cholesterol lowering drugs that target the mevalonate pathway, improve PDAC progression and survival (35,36). In this respect, our study shows that *PNLIPRP1* is involved in maintaining cholesterol homeostasis, with an enrichment of *Ldlr*, Srebp and mevalonate pathway genes in *Pnliprp1* KD. Furthermore, our genetic data further supports this hypothesis, as common and rare variants in the *PNLIPRP1* gene are associated with an increased LDL cholesterol.

In conclusion, we present evidence of an epigenetically-regulated decrease of *PNLIPRP1* with T2D milieu exposure in the pancreas contributing to dramatic changes in cellular fate of the exocrine pancreas. All these factors contribute towards the aggravated down-regulation of *PNLIPRP1*, inducing the trans-differentiation of acinar cells into duct-like cells, potentially making the cells particularly vulnerable to cancer initiation and PDAC progression (Figure 6).

**Figure 6.**
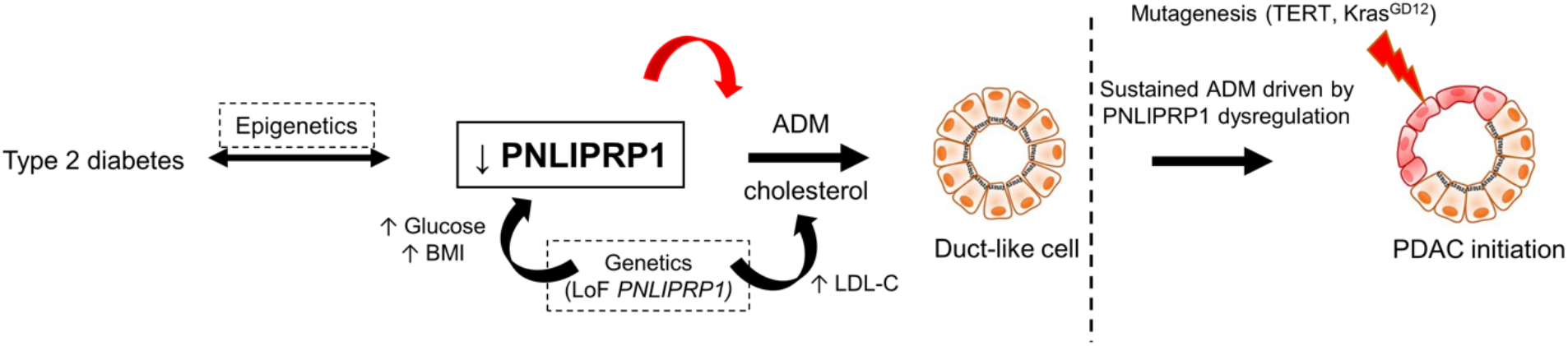
Proposed model of PNLIPRP1 role in T2D and ADM. We propose that the epigenetic dysregulation of *PNLIPRP1* aggravates hyperglycaemia, creating a feedback loop of *PNLIPRP1* downregulation. This down-regulation promotes cholesterol biosynthesis (supported by genetics) and ADM, creating a favourable environment of duct-like cells to progress into cancer initiation and PDAC.

Our study is unfortunately unique, as the exocrine pancreas is a tissue that is not usually conserved in biobanks, with the exception of GTEx, which has no phenotypic data of the tissue donors. However, we believe our epigenetic main result is supported by a body of functional evidence (not in human as no human acinar cell lines are available) and also genetic evidence. Despite all challenging issues working on the exocrine pancreas, further studies should decipher further the underlying molecular mechanisms linking T2D/obesity and future development of deadly PDAC, as a foundation for efficient prevention, early diagnosis and better treatments of PDAC.

## Supporting information

Manuscript Supplementary Tables

Manuscript Supplementary Figures

## Data Availability

All data produced in the present work are contained in the manuscript.

## Author contributions

The study was designed by AK and PF. LM, LoM, MS RB, and AL performed the wet lab experiments. PM, LM, AJ and SL provided clinical data. FP and JKC provided immunohistochemistry slides. BT, MD and SA performed RNA sequencing and bioinformatic analysis. AB and AA performed the analyses on rare variants and VP, JM and IP on common variants using the UK biobank. LN, MB and MC performed the statistical analyses for the EWAS and generated the associated figures and tables. AMS, MS and MI contributed data. AK, PF and LM interpreted the data and wrote the manuscript. All authors critically reviewed and edited the manuscript.

## Acknowledgements

We would like to thank Hutokshi Crouch for the help in generating the methylation data. Additional Files:

1) Additional File 1

All Supplementary Tables in the manuscript – Tables S1-S10

2) Additional File 2

All Supplementary Figures in the manuscript – Figures S1-S11

