## Supplementary material for "Epigenetics of *PNLIPRP1* in human pancreas reveals a molecular path between type 2 diabetes and pancreatic cancer": Manuscript Supplementary Figures

### 1. **Supplementary Methods**

### 1.1. GTEx to determine tissue expression of *PNLIPRP1*

The median gene-level transcript per million (TPM) by tissue from Genotype-Tissue Expression (GTEx) Portal (https://gtexportal.org/home/datasets/) was used to identify the expression pattern of *PNLIPRP1*.

### 1.1. RNA expression of *PNLIPRP1* and neighbouring genes in cohort

Prior to processing, the samples were stored in OCT at -80°C. To process the samples, the OCT was removed with a scalpel and the sample was cut into three pieces. The pieces were incubated in Trizol (15596-026; Thermofisher), 3% DTT and 5% B-Mercaptoethanol. During incubation, the samples were vortexed vigorously. Following incubation, the supernatant was collected and the RNA was extracted following the standard Trizol protocol (15596-026; Thermofisher). RNA was reverse transcribed using the High-capacity cDNA reverse Transcription Kit (Applied Biosystems; 4368814) following the manufacturer’s instructions. qPCRs were performed on the QuantStudio Pro 7 (Applied Biosystems). For cDNA quantification of selected targets, the SYBR green reagent mix was used (A25918; ThermoFisher). All primers used in this study are listed in Additional File 1: Table S2. Fold change was calculated using the ^∆∆^CT method (Livak and Schittgen, 2001). Two-tailed t-tests were performed using GraphPad Prism (GraphPad software Inc). Ten samples were processed (five controls and five T2D individuals), matched for age, sex and BMI (Additional File 1: Table S3).

### 1.2. Immunohistochemistry: protein expression of PNLIPRP1 in human tissues

Human pancreatic tissue sections, from control and T2D individuals, used in this study were donated by Inserm UMR1190 unit (University of Lille, France). We excluded any fibrotic process or pathological alterations in the pancreas tissues. Paraffin was removed from the slides with 100% xylene. Gradually decreasing concentrations of ethanol (100-50%) were used to rehydrate the tissue slides. Antigens were retrieved via incubation in sodium citrate buffer (pH 6). Non-specific site blocking was performed by incubating the slides in a 1x PBS, 0.01% triton, 5% goat serum solution for 30 min at room temperature (RT). Primary antibodies were incubated overnight at 4°C. Secondary antibodies were incubated for 1 hour at RT. Images were captured using Zeiss LSM 710 NLO confocal laser scanning microscope. Details of antibodies used are listed in Additional File 1: Table S4.

### 1.3. Functional characterisation

### 1.3.1. AR42J cell line – insulin responsive Akt

We verified whether the AR42J cells are insulin-responsive, by confirming the phosphorylation of Akt following insulin treatment. To do this, we plated and serum starved AR42J cells overnight and then washed with PBS and stimulated with or without 200 nM insulin for 1 hour sat 37 °C in 5% CO2. The total protein was harvested in RIPA buffer supplemented with protease and phosphatase inhibitors, as described below. The primary antibody (all used at 1:1000 dilution) used were anti pAKT (S473; Cell Signaling) and anti Akt (9272 - 1:5,000 dilution; Cell Signaling), and the secondary antibody used was goat pAb to Rb igG (Ab205718 - 1:20,000 dilution; Abcam).

### 1.3.2. Western blotting

Cells were lysed and protein harvested using a RIPA buffer (89900; ThermoFisher). Protein was quantified using the Pierce BCA Protein Assay Kit (23225; ThermoFisher) and separated on a 10% SDS-PAGE gel and transferred to a nitrocellulose membrane using the iBlot2 Gel Transfer Device (Life Technologies). Membranes were blocked with 5% non-fat dry milk or 5% BSA and incubated overnight at 4°C with desired antibody. Membranes were then incubated with an appropriate secondary antibody for 1 hour at room temperature (RT). Secondary antibody detection was performed using the LI-COR Biosciences imaging system. Protein expression was analysed with ImageJ.

### 1.3.3. MTS proliferation assay

AR42J cells (18,000 per well) were plated in a 96 well plate with 0% FBS medium and transfected with *Pnliprp1* siRNA or control siRNA, as described previously, in three biological replicates. Following a 72-hour incubation, the medium was removed and replaced with new 0% FBS medium and MTS proliferation assay (197010; Acbam) following the manufacturer’s instructions. After a one-hour incubation, 490 nm absorbance was measured with the iMark plate reader (Bio-Rad). Two-tailed t-tests were performed using GraphPad Prism (GraphPad software Inc).

### 1.3.4. Cholesterol content

AR42J cells were plated and transfected as described. After a 48 hours’ incubation period, the medium was removed and cells were washed with 100uL of PBS twice. Following this, the Cholesterol Ester-Glo Assay (J3190; Promega) was performed following the manufacturer’s instructions. The assay functions by producing NADH relative to cholesterol quantity. NADH then reduces proluciferin to luciferin. Luciferin can be quantified by measuring luminescence. Luminescence was quantified using the Glomax Microplate Luminometer (Promega). This experiment was performed in three biological replicates.

### 1.3.5. UK Biobank to identify rare and common variant associations

The equation of the MiST model is: $Y=\alpha X+\hat{\pi}GZ$ , where Y is the phenotype matrix (n x 1) for n individuals, X is the matrix of covariates (n x p) with p covariates, Z is a vector of q ones for the q variants and G is the genotype matrix (n x q) coded 0, 1 and 2 for AA, Aa, aa, where A is a major allele and a is a minor allele. In addition of MiST, we also extract the direct burden effect of the cluster $\hat{\pi}$ on the trait Y.

We utilised the Type 2 diabetes knowledge portal (https://t2d.hugeamp.org/) to assess whether common variants associated with T2D and related metabolic traits. The portal identifies gene-level phenotypic associations for *PNLIPRP1*, calculated from bottom-line genetic associations using the MAGMA (Multi-marker Analysis of GenoMic Annotation) method, with a significance MAGMA cut-off of p ≤ 2.5 x 10^-6^.

### 1.3.6. *PNLIPRP1* DNA variant association analyses for pancreatic cancer in UK Biobank and for T2D in publicly available summary statistics.

UK biobank dataset included 458,890 participants (1,425 cases; 457,465 controls), considering the individuals of Europeans ancestry only. We defined pancreatic cancer in UK Biobank using a combination of hospital admissions data, the tenth revision of the International Classification of Disease (ICD-10) codes and self-report data. Individuals with an ICD-10 (code C25) and who self-reported to have a pancreatic cancer diagnosis (code 1026) were set as cases, while individuals with no cancer diagnosis were set as controls.

In the *PNLIPRP1* gene region, we verified the associations with common and rare variant SNPs in two publicly available GWAS summary statistics datasets for T2D and PDAC, respectively. For analysis, we evaluated a DNA region window of five times the *PNLIPRP1* gene region. We used the latest T2D GWAS from Mahajan et al. (19), including 74,124 T2D cases and 824,006 controls of European ancestry.

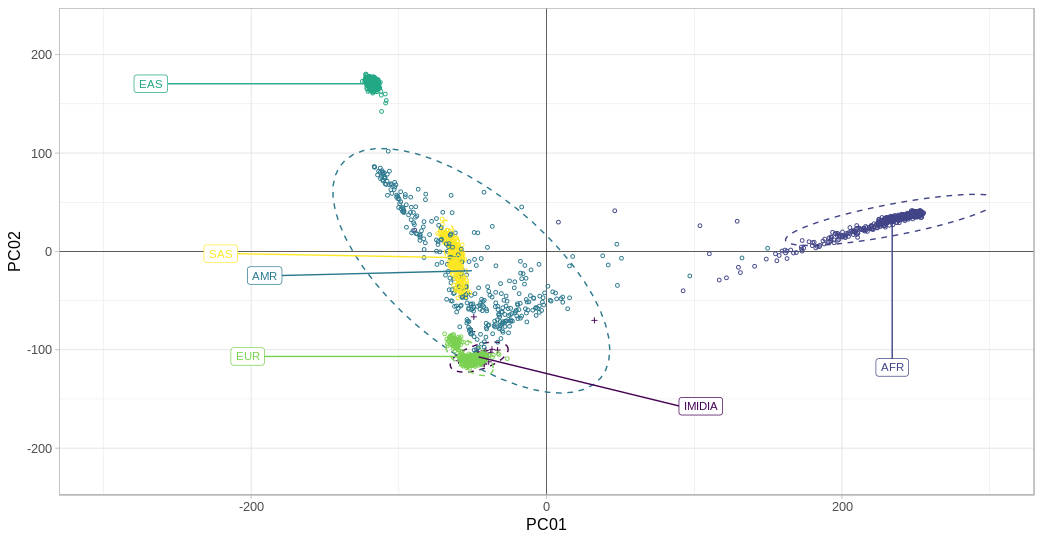

**Figure S1:** Ancestry clustering of samples from our samples (IMIDIA) confirmed European descent of subjects, compared to the 1,000 genomes.

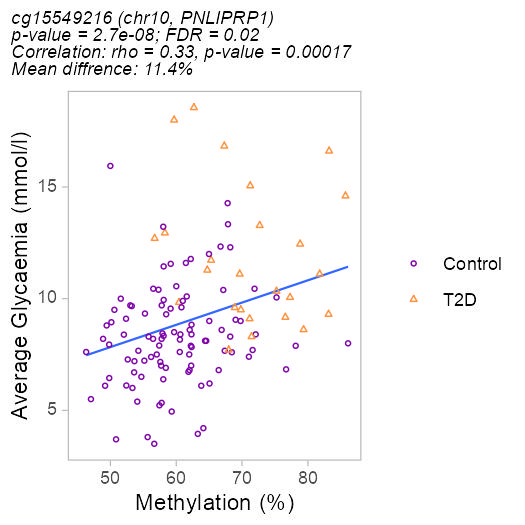

**Figure S2:** Scatterplot depicting the association between methylation level of cg15549216 and glucose levels in our cohort. Non-diabetic controls are shown in purple and T2D individuals are shown in orange. The correlation rho values were determined using a linear regression.

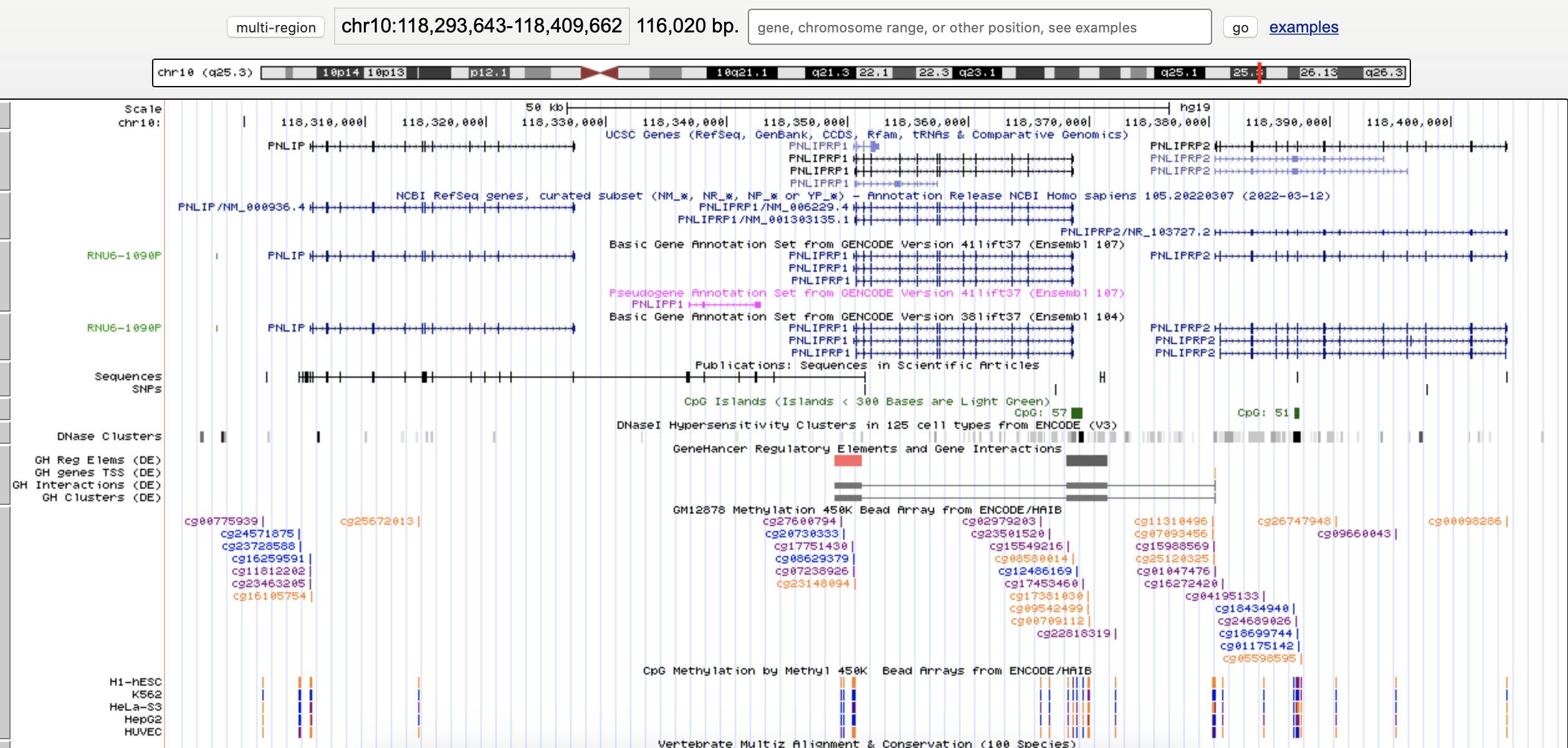

**Figure S3:** UCSC genome browser screenshot of the *PNLIPRP1* gene, surrounded by *PNLIP* and *PNLIPRP2.* The image illustrates the promoter (pink) and enhancer (grey) along with CpG islands (green).

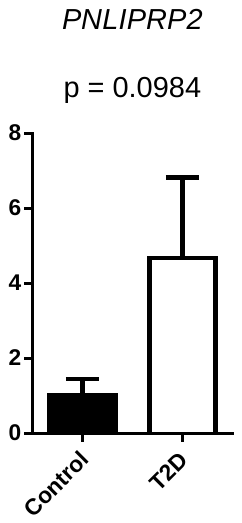

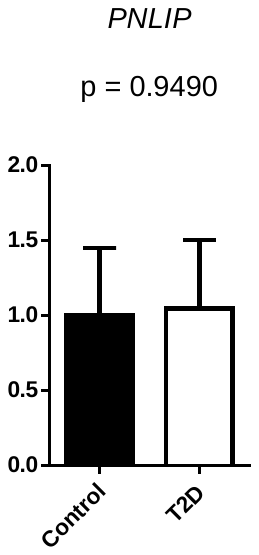

**Figure S4:** qPCR of *PNLIP* and *PNLIPRP2* from RNA of 6 controls and 5 T2D individuals, matched for age, sex and BMI. T-tests were performed to examine differences in expression between the control and T2D samples for each target gene. Error bars represent one standard error

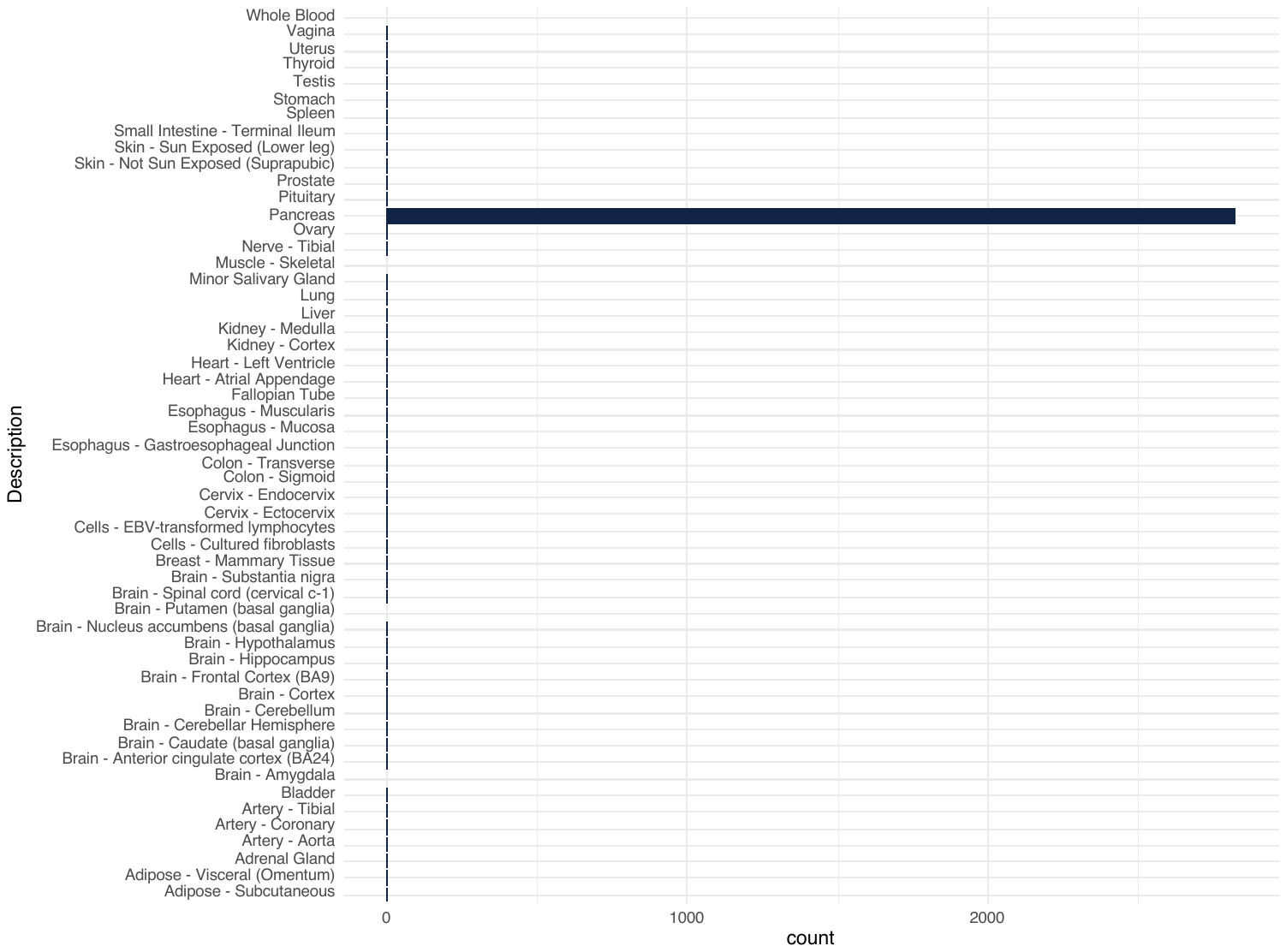

**Figure S5:** The expression of *PNLIPRP1* in the pancreas compared to all other tissues. Data was obtained from the GTEx public database. The count in the X-axis represents the TPM median.

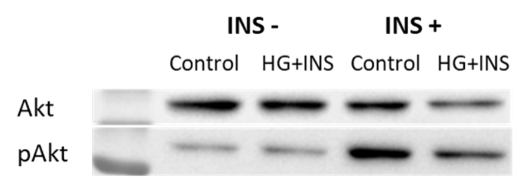

**Figure S6:** Western blot of Akt and phosphorylated Akt (pAkt) in AR42J stimulated with or without 200 nM insulin for 1 hour. The insulin treatment was performed following 48 hours of high glucose and insulin treatment.

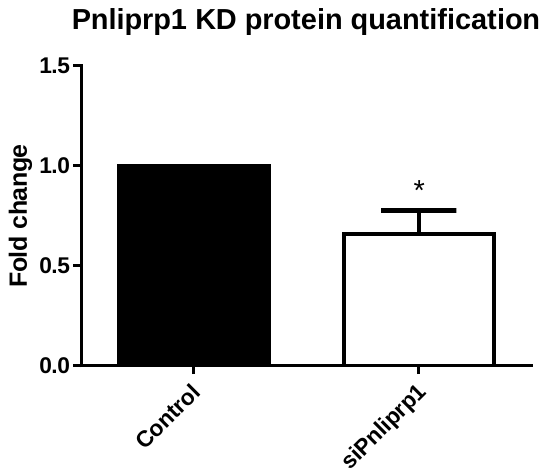

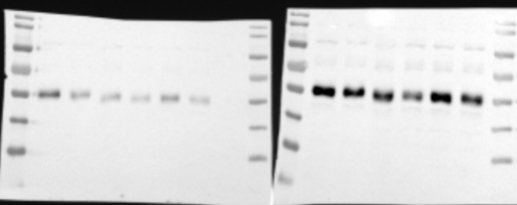

**Pnliprp1**

**B-actin**

siPnlirp1

siControl

siPnlirp1

siControl

siPnlirp1

siControl

siPnliprp1

siControl

siPnlirp1

siPnlirp1

siControl

siControl

**Figure S7:** Western blot of Pnliprp1 and housekeeping gene beta-actin following *Pnliprp1* KD (top). Quantification of the protein revealed a decreased expression of *Pnliprp1* in the KD compared to the non-targeting control. The difference was determined based on a two-tailed t-test. * p < 0.05.

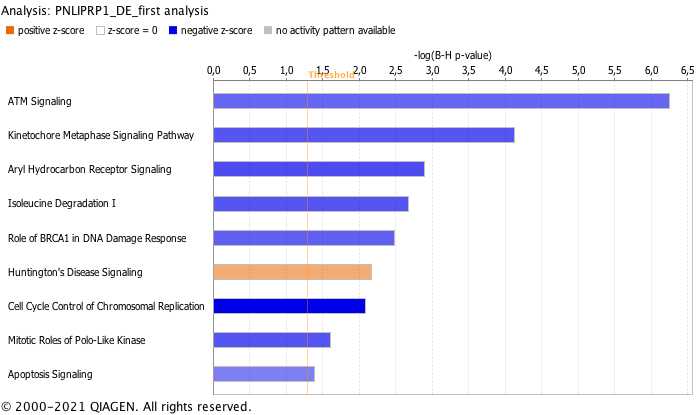

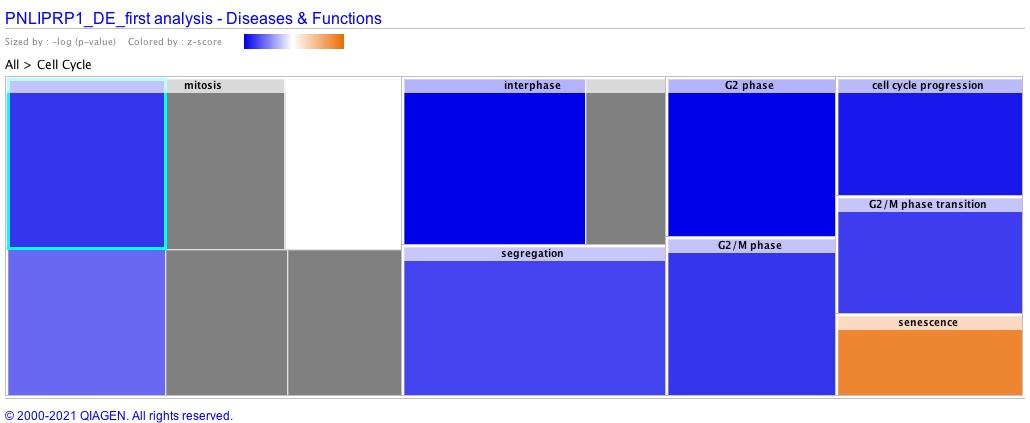

**Figure S8:** a) Ingenuity Pathway Analysis (IPA) database results of the dysregulated genes in *Pnliprp1* KD in AR42J. The pathways were organised according to significance using a Fisher’s exact test. A z-score of ±2 was used. A z-score of less than -2 indicates a significantly down-regulated pathway, presented in blue. b) IPA predicted the down-regulation of the cell cycle using our *Pnliprp1* KD RNA-seq data. Blue denotes a negative z-score (down-regulation) and orange a positive z-score (up-regulation) and in grey are the pathways where no activity pattern is predicted.

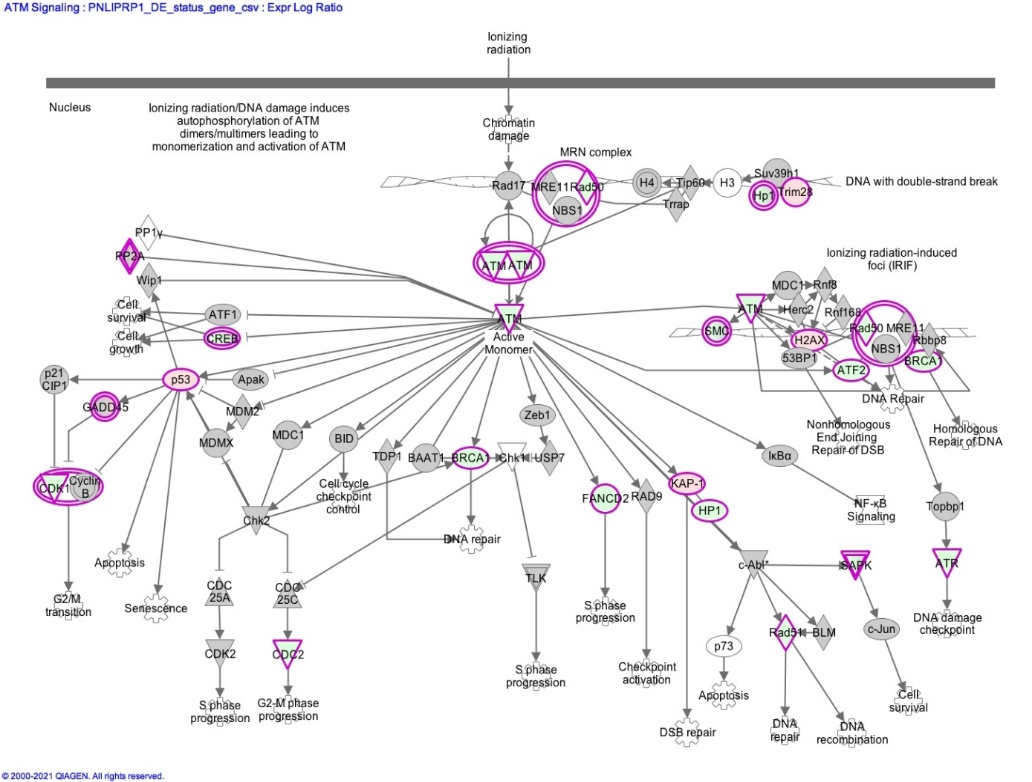

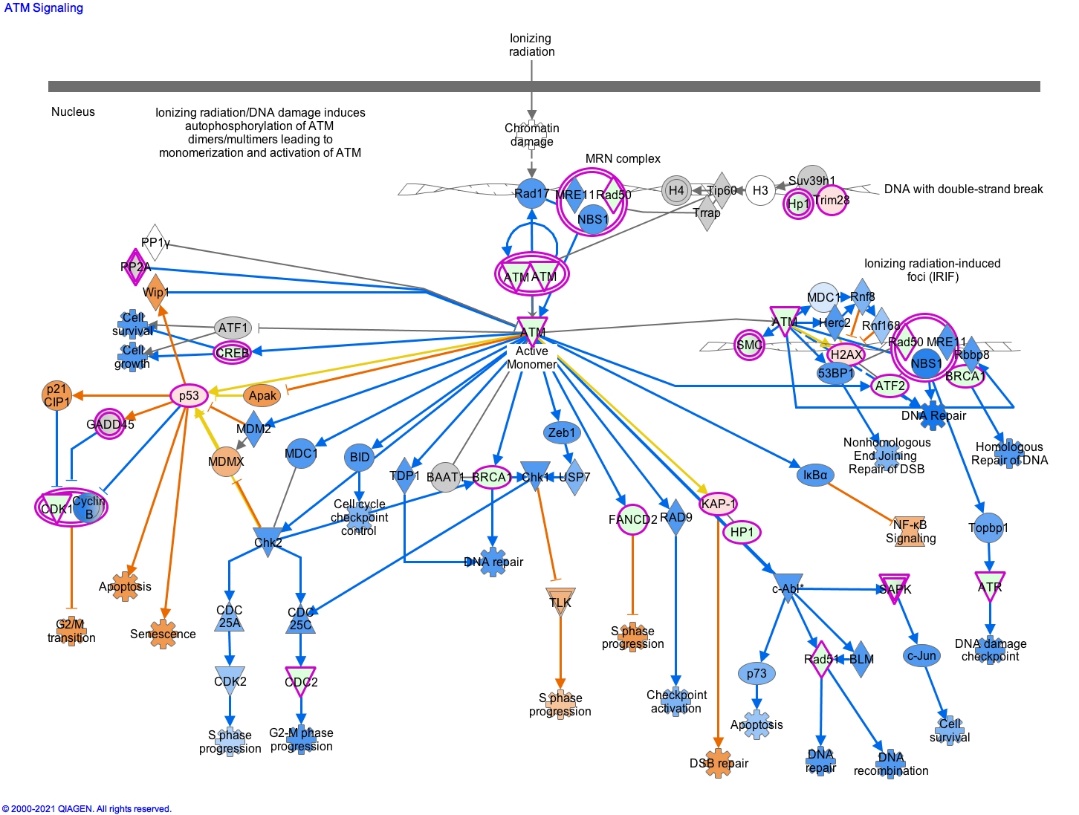

**Figure S9:** Overview of the ATM pathway. a) The ATM pathway was the most dysregulated pathway in our gene set as per IPA. The dysregulated genes identified in our *Pnliprp1* KD are shown in red (up regulated) or green (down-regulated). b) The same pathway, but with IPA predicting the up (orange) and downregulated (blue) downstream genes.

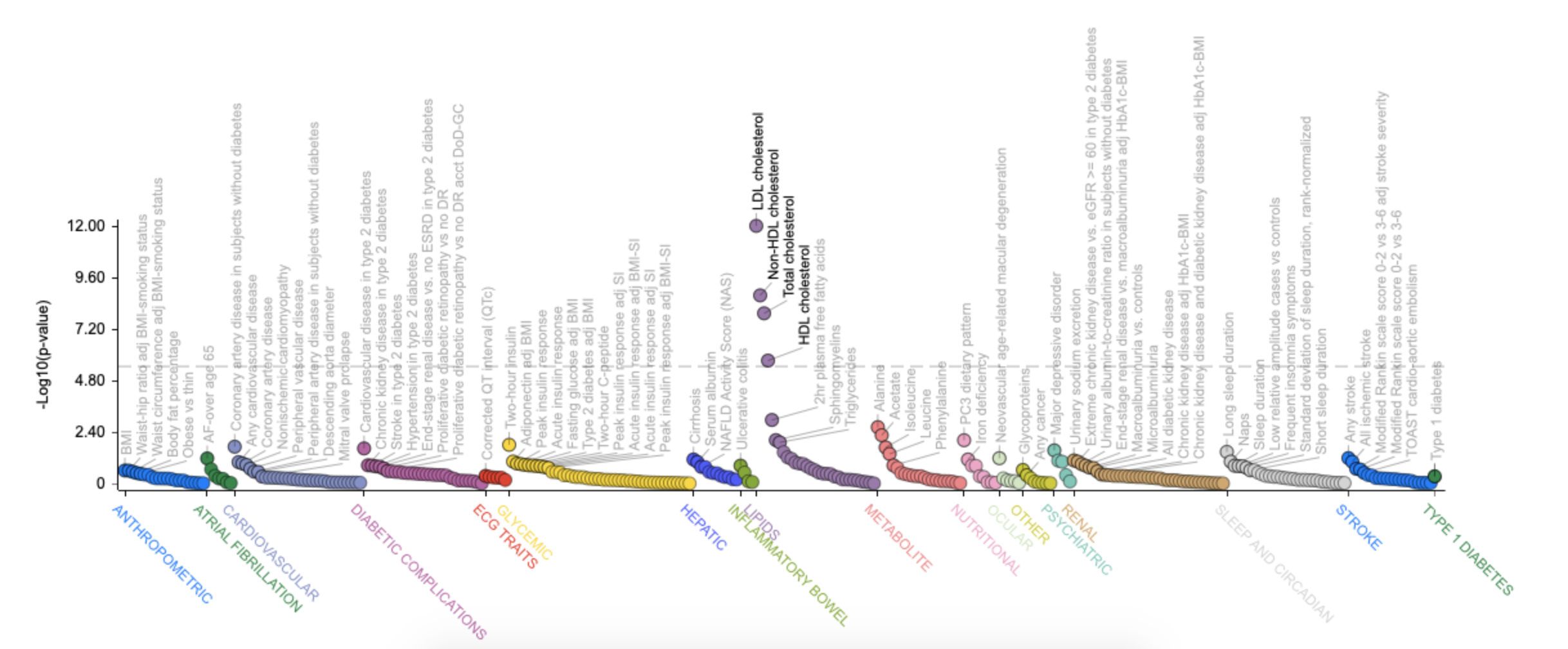

**Figure S10:** An overview of GWAS associations of SNPs in *PNLIPRP1* gene with type 2 diabetes associated traits, obtained from the Type 2 Diabetes Knowledge Portal. The plot shows gene-level phenotypic associations for *PNLIPRP1*, calculated from bottom-line genetic associations using the MAGMA (Multi-marker Analysis of GenoMic Annotation) method. The traits highlighted in black are significant following multiple testing.

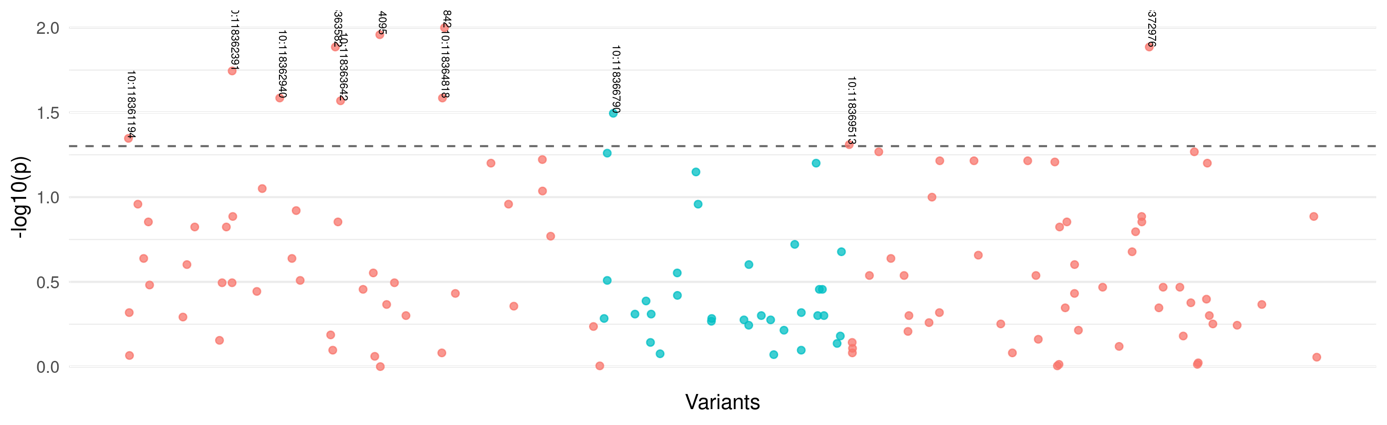

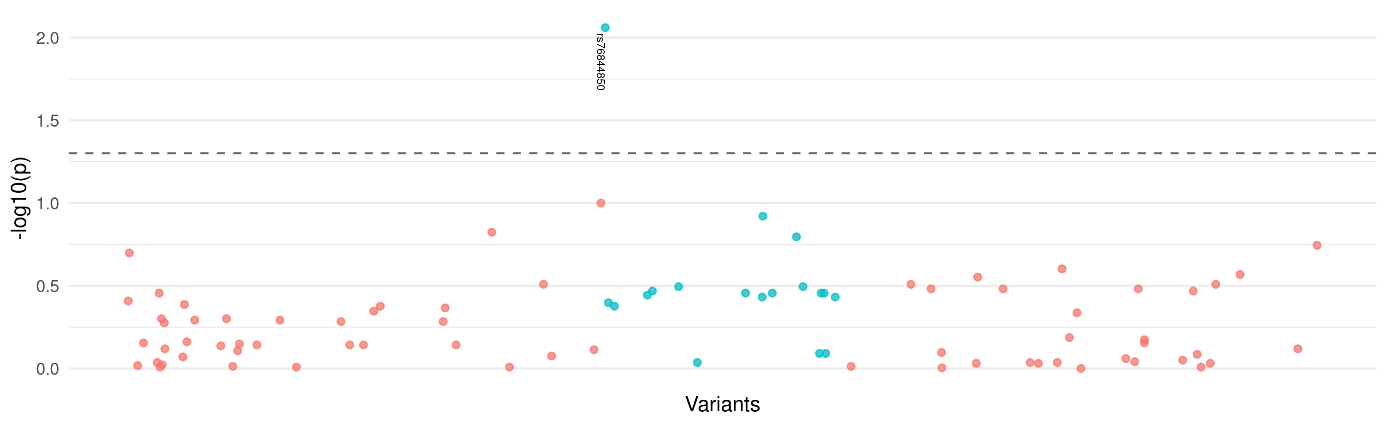
 **Figure S11:** GWAS associations of SNPs in *PNLIPRP1* gene with type 2 diabetes using the UK biobank. The dotted line represents nominal p-value. The SNPs indicated in blue represent the *PNLIPRP1* DMR region and in red the region surrounding the DMR.

**Figure S12:** GWAS associations of SNPs in *PNLIPRP1* gene with PDAC using the UK biobank. The dotted line represents nominal p-value. The SNPs indicated in blue represent the *PNLIPRP1* DMR region.
